# PINK1 Expression as a Prognostic Biomarker in Glioblastoma Multiforme: An Observational Multicenter Study

**DOI:** 10.64898/2026.04.03.26350127

**Authors:** Luis Alejandro García Rairán, Valentina Corpus Gutiérrez, Maria A. Del Castillo, William Riveros Castillo, Javier Saavedra Gerena, Andres David Turizo Smith, Jaime Arias Guatibonza

**Affiliations:** Departamento De Neurocirugía, Facultad de Medicina, Universidad Nacional de Colombia, Bogotá 111221, Colombia; Center for Research and Training in Neurosurgery (CIEN), Samaritana University Hospital, Bogotá, Colombia; Grupo de investigación INFARMA-HUS, Hospital Universitario de la Samaritana, Cra 8 N.° 0-29 sur, Bogotá 111411, Colombia; Grupo de investigación en Fitoquímica y Farmacognosia (GIFFUN), Universidad Nacional de Colombia,Bogotá 111221, Colombia

**Keywords:** Glioblastoma (GBM), PINK1, Prognostic marker, Neurosurgery, Neurological oncology, Overall Survival (OS), Progression Free Survival (PFS), Quality Of Life (QoL)

## Abstract

**Introduction:** Glioblastoma multiforme (GBM) remains the most lethal primary brain tumor with median survival of 14-15 months. Current prognostic markers inadequately stratify patient outcomes. PINK1 (PTEN-induced putative kinase 1), a mitochondrial kinase regulating mitophagy and cellular stress responses, has emerged as a promising prognostic candidate. Our preliminary analysis of 20 GBM cases demonstrated significant PINK1 expression with correlation to aggressive phenotypes (Turizo Smith et al., 2025). This multicenter study aims to validate PINK1 as a prognostic biomarker for survival and functional outcomes in a Latin American cohort.

**Methods and analysis:** PINK1-GBM Colombia is a multicenter, observational cohort study across four tertiary hospitals in Bogotá, Colombia (Hospital de Kennedy, Hospital El Tunal, Hospital Santa Clara and Hospital Universitario de la Samaritana). We will enroll at least 26-50 adults (≥18 years) with newly diagnosed IDH-wild type GBM undergoing surgical resection. PINK1 expression will be quantified by immunohistochemistry (IHC) on formalin-fixed paraffin embedded (FFPE) tissue using standardized protocols. Primary outcomes: overall survival (OS) and progression-free survival (PFS). Secondary outcomes: functional status trajectories (KPS/ECOG), Follow-up extends 24 months with clinical, imaging (RANO 2.0), and telephone assessments. Survival analyses will employ Kaplan-Meier methods, log-rank tests, and Cox proportional hazards models adjusted for established prognostic factors.

**Ethics and dissemination:** Approved by Universidad Nacional de Colombia Ethics Committee (Acta 001, February 5, 2026; Ref: 2.FM.1.002-CE-002-26), Subred Sur Occidente (P-AP-19-2025, July 11, 2025), and Subred Centro Oriente (CEI 067/2025, October 24, 2025). Conducted per Declaration of Helsinki and Colombian Resolution 8430/1993. Results will be disseminated via peer-reviewed publication, international conferences, and thesis submission.

**Trial registration:** [To be registered at ClinicalTrials.gov]

## Introduction

Glioblastoma multiforme (GBM, WHO CNS grade 4) represents 48.6% of all malignant primary central nervous system tumors, with an annual incidence of 3.19 per 100,000 worldwide [1]. Despite maximal safe resection, concurrent chemoradiation with temozolomide (TMZ), and adjuvant TMZ, median overall survival remains 14.6 months, with 5-year survival below 5% [2,3]. The inevitable recurrence and therapeutic resistance underscore the critical need for robust prognostic biomarkers to stratify patients and guide personalized treatment strategies.

Current molecular markers include MGMT (O6-methylguanine-DNA methyltransferase) promoter methylation, which predicts TMZ response, and IDH (isocitrate dehydrogenase) mutation status, which indicates favorable prognosis [4,5]. However, these markers incompletely explain outcome heterogeneity, particularly in IDH-wild type GBM, which constitutes approximately 90% of primary GBMs [6]. Novel biomarkers reflecting tumor biology and host response are urgently needed.

### PINK1: Biological Rationale and Preclinical Evidence

PINK1 (PTEN-induced putative kinase 1) is a mitochondrial serine/threonine kinase encoded on chromosome 1p36.12. Under physiological conditions, PINK1 is imported into mitochondria and degraded by presenilin-associated rhomboid-like protease (PARL). Upon mitochondrial membrane depolarization, PINK1 stabilizes on the outer membrane, recruiting the E3 ubiquitin ligase Parkin to initiate mitophagy—selective autophagy of dysfunctional mitochondria [7,8].

Beyond mitophagy, PINK1 regulates multiple cellular processes relevant to cancer biology:

- Apoptosis inhibition: PINK1 phosphorylates Bcl-xL at serine 62, preventing its pro-apoptotic cleavage and stabilizing mitochondrial membranes [9,10]
- Cell cycle regulation: PINK1 deficiency causes G2/M arrest and reduced proliferation through p53-dependent mechanisms [11]
- Oxidative stress response: PINK1 loss increases reactive oxygen species (ROS) production, potentially sensitizing cells to therapy-induced damage [12,13]
- Immune modulation: PINK1 affects microglial activation and neuroinflammation through regulation of mitochondrial antigen presentation [14]

### Context-Dependent Function of PINK1 in Cancer

PINK1 exhibits a context-dependent functional duality that varies according to cancer type, cellular microenvironment, and specific molecular interactions. In glioblastoma, PINK1 functions predominantly as a tumor suppressor, where its epigenetic silencing or deletion of the 1p36 locus correlates with more aggressive phenotypes and reduced survival [13]. Agnihotri et al. demonstrated that PINK1 is a negative regulator of growth and the Warburg effect in glioblastoma, with PINK1-negative gliomas showing significantly reduced survival in patients treated with temozolomide and radiotherapy [13]. The tumor suppressor function extends to lung cancer, where Lin et al. showed that PINK1 blocks EGFR-driven tumorigenesis through physical interaction via PINK1-CTD, preventing EGFR dimerization and subsequent oncogenic signaling [15]. In pancreatic cancer, PINK1 suppresses tumorigenesis through control of mitochondrial iron-mediated immunometabolism, where PINK1-mediated mitophagy degrades iron importers to limit mitochondrial iron accumulation [16]. However, PINK1 can also exhibit oncogenic potential depending on the cellular context. In lung adenocarcinoma, high PINK1 expression has been associated with chemoresistance and poor treatment response [17]. Similarly, in esophageal squamous cell carcinoma, high PINK1 expression correlates with poor prognosis, suggesting that PINK1 may promote tumor survival under specific metabolic conditions or therapeutic pressures [17]. This functional plasticity suggests that PINK1’s role could be modified by the metabolic state of the tumor, with its tumor suppressor functions predominating in contexts of mitochondrial stress and EGFR-driven proliferation, while potentially supporting tumor survival in hypoxic or nutrient-deprived conditions where mitophagy provides essential metabolic adaptation.

### PINK1 in Glioma: Clinical Evidence

The clinical relevance of PINK1 in glioma was first systematically investigated by Agnihotri et al. (2016), who demonstrated that PINK1-negative gliomas had significantly reduced survival in patients treated with TMZ and radiotherapy [13]. Key findings include:

- PINK1 mRNA expression decreases with tumor grade, with lowest levels in GBM
- The 1p36 deletion, common in gliomas (50-70% of cases), directly targets the PINK1 locus
- PINK1 loss correlates with increased proliferation markers and reduced apoptosis
- In medulloblastoma, PINK1 loss occurs across all molecular subgroups [16].

More recently, the interaction between PINK1 and the epidermal growth factor receptor (EGFR) pathway has emerged as a critical mechanism underlying PINK1-mediated tumor suppression in cancer [15]. This interaction is particularly relevant given that EGFR amplification and the +7/-10 chromosomal signature represent defining molecular features of IDH-wildtype glioblastoma.

The epidermal growth factor receptor (EGFR) represents one of the most relevant molecular targets in glioblastoma multiforme (GBM), with amplification or mutation present in approximately 40-50% of cases, particularly the EGFRvIII variant that lacks the ligand-binding extracellular domain [3]. Recent evidence has elucidated a direct molecular interaction between PINK1 and EGFR: PINK1 physically interacts with EGFR via its C-terminal domain (PINK1-CTD) and the EGFR tyrosine kinase domain, constituting an endogenous steric hindrance to receptor dimerization that inhibits EGFR-mediated tumorigenesis [15]. In lung cancer models, PINK1 depletion promoted EGFR dimerization, receptor activation, and downstream signaling, whereas PINK1 overexpression or PINK1-CTD administration suppressed these oncogenic processes [15]. This mechanism is particularly relevant in the context of the +7/-10 chromosomal signature, which is present in 60-70% of GBM cases and frequently co-occurs with EGFR amplification [18]. According to Rodriguez-Moreno et al., the +7/-10 cytogenetic alteration is the most frequent chromosomal abnormality in GBM and serves as a defining diagnostic criterion for isocitrate dehydrogenase wild-type (IDHwt) glioblastoma under the 2021 WHO classification [18]. The signature arises early in tumorigenesis, often years before clinical diagnosis, and establishes a common evolutionary path across IDHwt GBM [19]. Nair et al. proposed that the frequent co-occurrence of chromosome 7 gain and chromosome 10 loss is driven by a synthetic rescue mechanism: loss of chromosome 10 removes key tumor suppressor genes including PTEN, ANXA7, and KLF6, which while favoring tumor progression also disrupt essential cellular processes and impose a significant fitness cost [20]. To survive this, tumor cells upregulate compensatory genes on chromosome 7, including EGFR, which can functionally rescue PTEN loss, and MET, which compensates for ANXA7 loss [20]. This phenomenon reflects an adaptive evolutionary response whereby chromosome 7 gain mitigates the detrimental effects of chromosome 10 loss through a distributed compensatory network [18,20]. EGFR amplification, typically mapped to 7p11.2, tends to appear after complete chromosome 7 gain and is associated with increased pathway activation and tumor proliferation [18]. The coexistence of +7/-10, EGFR amplification, and TERT promoter mutation has shown specificity of up to 99.4% for IDHwt GBM and is absent in non-GBM entities [18]. In this study, the evaluation of EGFR by immunohistochemistry will allow exploration of potential correlations between PINK1 status and EGFR expression, which could identify patient subgroups with distinct molecular vulnerabilities and potentially guide combined therapeutic strategies targeting both pathways.

Subsequent studies have confirmed that mitophagy dysregulation, particularly through PINK1/Parkin pathway alterations, contributes to TMZ resistance in glioma cells [17,21]. The SUMOylation status of PINK1, regulated by SENP6, appears critical for determining TMZ sensitivity [22].

### Rationale for This Study

Despite compelling preclinical and translational evidence, clinical validation of PINK1 as a prognostic biomarker in GBM is lacking. Existing studies are retrospective, predominantly from North American and European cohorts, and lack standardized assessment protocols. This study addresses these gaps by:

- Prospective and retrospective design with predefined endpoints and standardized IHC methodology
- Focus on an understudied population (Latin American patients)
- Comprehensive functional assessment alongside survival outcomes
- Integration with established prognostic factors (EGFR, IDH, clinical variables)

Hypothesis: Alterations in PINK1 expression in GBM tumor tissue is associated with shorter overall survival, shorter progression-free survival, and worse functional status, independent of established prognostic factors.

## Methods And Analysis

### Study Design

Multicenter, observational cohort study conducted according to SPIRIT (Standard Protocol Items: Recommendations for Interventional Trials) guidelines [23]. The study protocol was developed using the SPIRIT checklist (provided as online supplementary material).

### Study Setting

Four tertiary care public hospitals in Bogotá, Colombia:

1. Hospital de Kennedy
2. Hospital El Tunal
3. Hospital Santa Clara
4. Hospital de la Samaritana (HUS), ethics approval pending

These institutions provide neurosurgical oncology attention to the majority of GBM patients in Colombia’s capital region, serving a diverse socioeconomic population.

### Eligibility Criteria

#### Inclusion

- Age ≥18 and ≤80 years
- Histologically confirmed, newly diagnosed IDH-wild type GBM (WHO 2021 classification) [24]
- Undergoing surgical resection (maximal safe resection or biopsy)
- Available FFPE tissue for IHC analysis
- Karnofsky Performance Status (KPS) ≥70 at enrollment
- Signed informed consent for research participation

#### Exclusion

- Prior cranial radiation or chemotherapy for any CNS tumor
- Recurrent GBM
- IDH-mutant glioma (per WHO 2021 criteria)
- Other concurrent malignancy (except adequately treated basal cell carcinoma, squamous cell carcinoma, or carcinoma in situ)
- Severe comorbidities limiting life expectancy <12 weeks
- Inability to complete follow-up assessments

### Sample Size Estimation

We anticipate 26-60 evaluable patients based on expected accrual across participating centers over 30 months. With 60 patients, assuming 30% PINK1-negative (based on Turizo-Smith et al. [25,26] and using Schoenfeld’s formula for Cox proportional hazards models, we will have 80% power (α=0.05, two-sided) to detect a hazard ratio of 2.0 for survival difference between PINK1-positive and PINK1-negative groups, assuming median survival of 18 vs. 10 months and 24-month follow-up [27].

### Recruitment and Consent

Patients with radiological suspicion of GBM will be identified through neurosurgical consultation based on contrast-enhanced magnetic resonance imaging (MRI) demonstrating: (i) T1-weighted hypointense lesion with irregular ring-enhancement following gadolinium administration; (ii) central necrosis; (iii) perilesional T2/FLAIR hyperintensity representing vasogenic edema; (iv) mass effect with midline shift ≥5mm or ventricular compression; and (v) restricted diffusion on diffusion-weighted imaging (DWI) suggesting high cellularity. Advanced MRI sequences including perfusion-weighted imaging (PWI) and magnetic resonance spectroscopy (MRS) will be obtained when available per institutional protocols, though their acquisition is not mandatory for study eligibility given variable availability across participating centers. Following informed consent for clinically indicated surgery, separate research consent will be obtained for:

1. Collection of additional tumor tissue for research
2. Review of medical records and imaging
3. Longitudinal follow-up assessments
4. Future use of residual tissue for related research

Enrollment will occur August 2025–December 2027. Patients may withdraw consent at any time without affecting clinical care.

### Tissue Collection and Processing

| Stage | Procedure | Standard | Responsible |
| --- | --- | --- | --- |
| Intraoperative | Additional tumor sampling | ≥3-5 cm<sup>3</sup> from viable tumor margin; avoidance of necrotic center | Neurosurgeon (L.A. García Rairán) |
| Immediate handling | Placement in sterile container with 10% neutral buffered formalin | Volume ratio 1:10 (tissue:formalin) | Surgical team |
| Fixation | 24-48 hours at room temperature | Agitation at 4 hours; tissue thickness <5mm | Hospital pathology |
| Transport | Coded container (Study ID only) to processing lab | Temperature 15-25°C; chain of custody documentation | Research coordinator |
| Processing | FFPE block preparation | Automated tissue processor; paraffin embedding within 7 days | Dr. L.R. Rengifo |
| Sectioning | 4µm sections for IHC; additional unstained slides for future use | Positive charged slides; ribbon technique | Histotechnologist |
| IHC staining | PINK1 immunohistochemistry | Standardized protocol (see below) | Immunotech Laboratories S.A.S. |
| Storage | FFPE blocks and stained slides | 15-25°C, dark, dry; minimum 5 years per Colombian regulations | Immunotech Laboratories S.A.S. |

### Immunohistochemistry Protocol

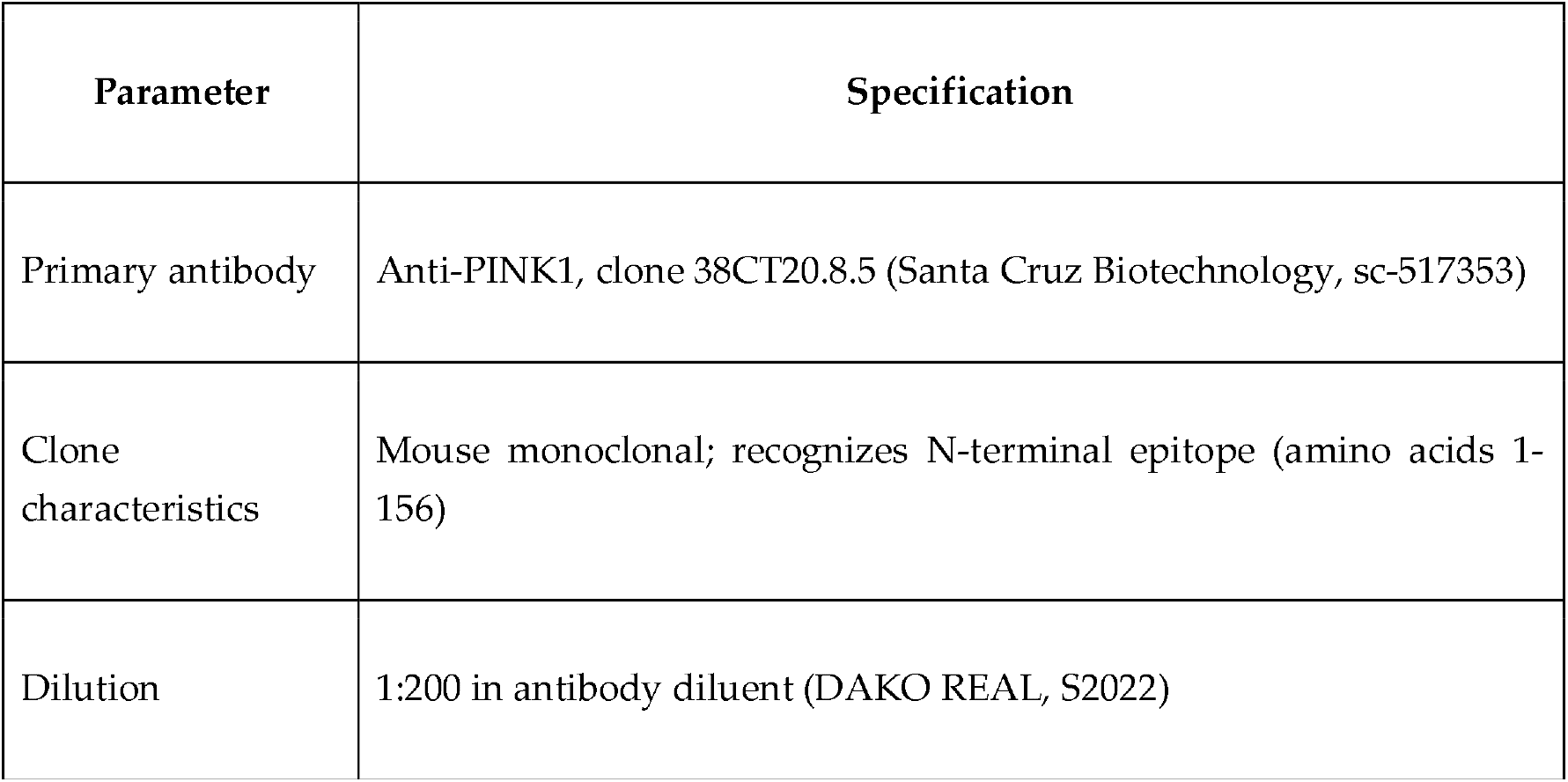
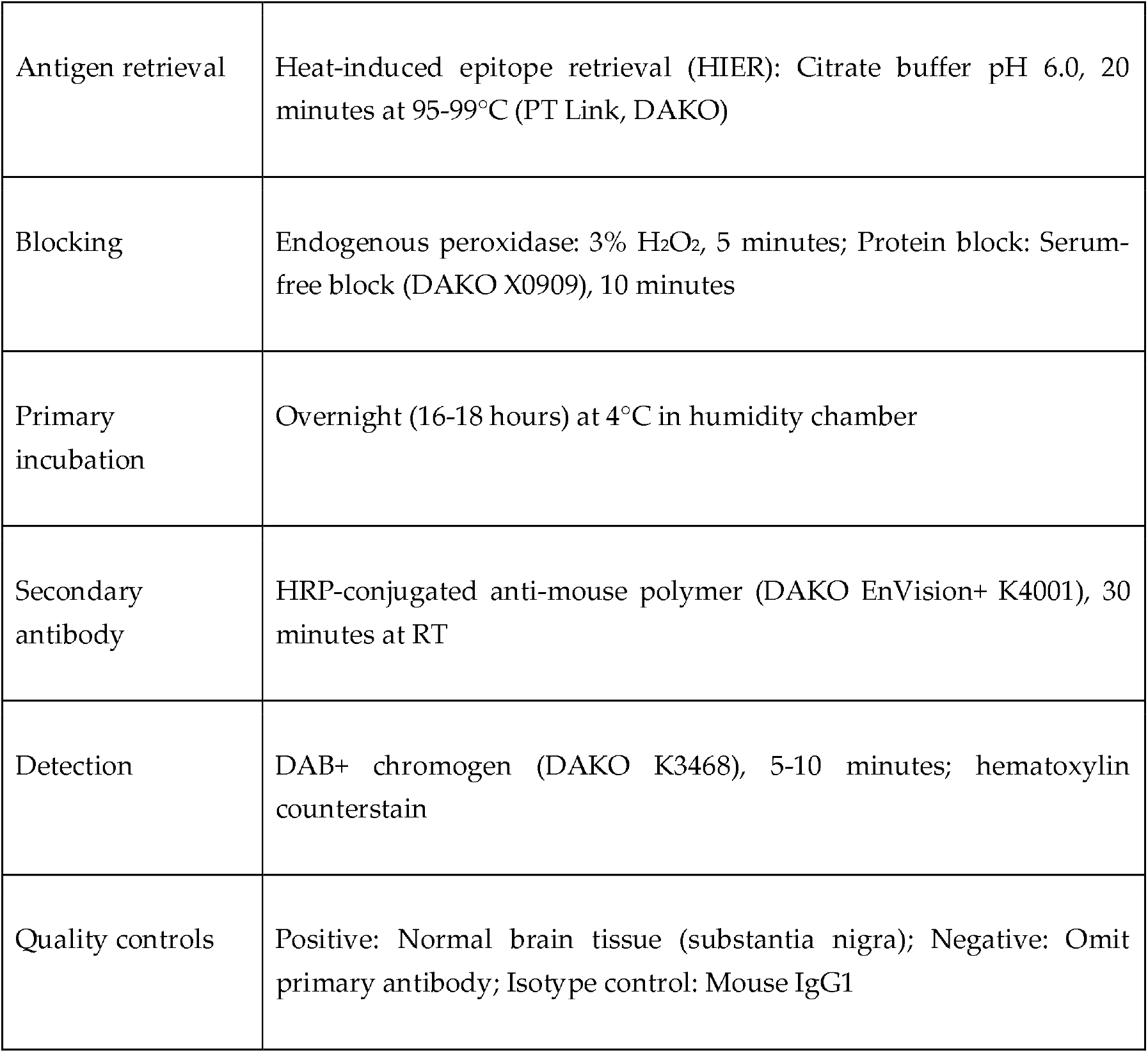

This protocol is adapted from published methodologies [28,29].

### PINK1 Scoring System

A single pathologist blinded to clinical outcomes will evaluate all samples:

1. Qualitative assessment: Present (any detectable staining) vs. Absent (no staining)
2. Semiquantitative H-score (if present):
  - Intensity: 0 (none), 1 (weak), 2 (moderate), 3 (strong)
  - Percentage positive cells: 0%, 1-5%, 6-25%, 26-50%, 51-75%, 76-100%
  - H-score = Σ(Intensity × Percentage), range 0-300

### Study Assessments and Follow-up Schedule

| Timepoint | Window | Clinical | Imaging | EOT Assessment | Functional | Biomarker |
| --- | --- | --- | --- | --- | --- | --- |
| Baseline | Preoperative | History, neuro exam, comorbidities | MRI brain (contrast-enhanced, perfusion, spectroscopy) | — | KPS, ECOG | — |
| Postoperative | 24-72 hours | Surgical complications, pathology | MRI (early post-op, 72 hours) | Validated EOT per RANO criteria | KPS | IDH IHC, PINK1, EGFR |
| Week 2-4 | ±7 days | Wound check, treatment planning | — | — | ECOG (telephone) | — |
| Month 3 | ±14 days | Clinical evaluation | MRI (RANO 2.0) | — | KPS, ECOG | — |
| Month 6 | ±14 days | Clinical evaluation | MRI (RANO 2.0) | — | KPS, ECOG | — |
| Month 9 | ±14 days | Telephone assessment | — | — | ECOG | — |
| Month 12 | ±14 days | Clinical evaluation | MRI (RANO 2.0) |  | KPS, ECOG | — |
| Month 18 | ±14 days | Telephone assessment | MRI (if indicated) | — | KPS, ECOG | — |
| Month 24/End | ±30 days | Final assessment | MRI (if indicated) | — | KPS, ECOG | — |

### Response Assessment

Radiological response will be assessed per Response Assessment in Neuro-Oncology (RANO) 2.0 criteria [30]:

- Complete response: Disappearance of all enhancing disease
- Partial response: ≥50% decrease in sum of perpendicular diameters
- Stable disease: <50% decrease and <25% increase
- Progressive disease: ≥25% increase or new lesion

Pseudoprogression will be distinguished from true progression using advanced MRI sequences (perfusion, spectroscopy) and clinical correlation, per RANO 2.0 guidelines.

### Outcomes

#### Primary outcomes

1. Overall survival (OS): Time from surgery to death from any cause
2. Progression-free survival (PFS): Time from surgery to radiological progression (RANO 2.0) or death, whichever occurs first

#### Secondary outcomes

1. Functional status trajectory (KPS and ECOG scores over 24 months)
2. Association between PINK1 expression and 6-month PFS
3. Correlation between PINK1 H-score (continuous) and survival outcomes
4. Time to functional decline (defined as KPS decrease ≥20 points or ECOG increase ≥2 levels)

#### Exploratory outcomes

1. PINK1 expression correlation with tumor location, size, and molecular profile
2. Patterns of failure (local vs. distant/multifocal) by PINK1 status
3. Cost-effectiveness of PINK1 testing for prognostic stratification

### Statistical Analysis Plan

#### Descriptive statistics

- Continuous variables: Mean (SD) or median (IQR) depending on distribution
- Categorical variables: Frequencies and percentages
- Baseline characteristics stratified by PINK1 status

#### Primary analysis

- Kaplan-Meier curves for OS and PFS by PINK1 status (present/absent)
- Log-rank test for between-group differences
- Cox proportional hazards models for PINK1 effect on OS/PFS, adjusted for: Age (continuous)

#### Sex (male/female)

- Preoperative KPS (≥80 vs. 70-79)
- Extent of resection (gross total vs. subtotal/biopsy)
- Tumor location (eloquent vs. non-eloquent area)
- Treatment compliance (completed standard chemoradiation vs. not)

#### Secondary analyses

- Linear mixed models for functional status trajectories
- Logistic regression for 6-month progression
- Subgroup analyses: EGFR status, age <65 vs. ≥65, extent of resection
- Sensitivity analyses: Multiple imputation for missing data; competing risks for non-cancer deaths; per-protocol vs. intention-to-treat

Software: R version 4.3+ (survival, survminer, lme4 packages) and GraphPad Prism [31,32]

### Missing Data and Loss to Follow-up

- Multiple imputation by chained equations (MICE) for missing covariates
- Inverse probability weighting for attrition
- Sensitivity analysis: Best-case/worst-case scenarios for missing outcomes

### Bias Mitigation Strategies

| **Potential Bias** | **Mitigation Strategy** |
| --- | --- |
| Selection bias | Consecutive enrollment using predefined eligibility criteria; screening log of all eligible patients; inclusion regardless of prognosis; multicenter design. |
| Information bias | Standardized case report forms; trained research personnel; central imaging review when feasible. |
| Measurement bias | Single blinded pathologist; validated IHC protocol with positive and negative controls; random 10% sample re-scoring for inter-assessment quality control. |
| Attrition bias | Multiple contact methods (telephone, email, family contact); telephone assessments permitted; documentation and reporting of reasons for loss to follow-up. |
| Confounding | Prespecified covariates for multivariable adjustment; propensity score sensitivity analyses; E-value estimation for unmeasured confounding [33]. |

### Ethics And Dissemination

This study involves minimal risk as tissue collection occurs during clinically indicated surgery, with no additional procedures or interventions. The study complies with:

- Declaration of Helsinki (latest revision, 2013) [34]
- Colombian Resolution 8430/1993 (research ethics regulations) [35]
- Law 1581/2012 (personal data protection)
- Good Clinical Practice guidelines (ICH-GCP E6 R2)

#### Ethics Approvals Obtained

Ethics Approvals Obtained:
| Institution | Reference | Date | Act Number | Status |
| --- | --- | --- | --- | --- |
| Universidad Nacional de Colombia | 2.FM.1.002-CE-002-26 | February 5, 2026 | Acta 001 | Active |
| Subred Integrada de Servicios de Salud Sur Occidente | P-AP-19-2025 | July 11, 2025 | Acta 12 | Active |
| Subred Integrada de Servicios de Salud Centro Oriente | CEI 067/2025 | October 24, 2025 | Acta 473 | Active |
| Subred Integrada de Servicios de Salud Sur | 20255000-030118-1 | November 7, 2025 | Acta 320 | Active |
| Hospital de la Samaritana | — | Submitted April 1, 2026 | — | Pending |

Approvals pending from Hospital Universitario Nacional, Clínica Palermo and Hospital de la Samaritana. The study will not commence at these sites until institutional approval is obtained.

### Informed Consent

- The informed consent process includes:
- Explanation of study purpose, procedures, risks, and benefits
- Clarification that participation is voluntary and does not affect clinical care
- Separate consent for tissue banking and future use
- Right to withdraw without penalty
- Contact information for questions or concerns

Consent forms are available in Spanish (see online supplementary material).

### Confidentiality and Data Protection

- All data coded with unique study identifiers; key linking codes to identifiers stored separately and securely
- Electronic data stored on encrypted, password-protected servers
- Paper records in locked cabinets with restricted access
- No identifying information in publications or presentations

### Return of Results

Individual PINK1 results will be shared with participants upon request through their treating physician, with clear explanation that:

- Results are investigational and not clinically validated
- No treatment decisions should be based on PINK1 status
- Results may be included in future clinical validation studies

### Dissemination Plan

1. Protocol publication: BMJ Open or Trials
2. Primary results: Journal of Neuro-Oncology, Neuro-Oncology, or Cancers
3. Secondary analyses: Specialty journals (Neurosurgery, Frontiers in Oncology)
4. Conference presentations:
  - Colombian Association of Neurosurgery (ACNC)
  - Society for Neuro-Oncology (SNO) Annual Meeting
  - American Association for Cancer Research (AACR)
  - European Association of Neuro-Oncology (EANO)
5. Thesis: Neurosurgery specialty degree, Universidad Nacional de Colombia
6. Data sharing: Deidentified dataset and statistical code available upon reasonable request to corresponding author, per FAIR principles [36]

### Authorship

- Authorship will follow ICMJE criteria [37]:
- Substantial contributions to conception/design, data acquisition, analysis, or interpretation
- Drafting or critical revision of intellectual content
- Final approval of published version
- Agreement to be accountable for all aspects of the work

### Strengths And Limitations

#### Strengths

- Prospective and retrospective design with predefined endpoints and analysis plan, reducing risk of bias
- Multicenter recruitment enhancing generalizability to Colombian/Latin American populations
- Standardized IHC protocol with blinded assessment
- Comprehensive functional status assessment alongside survival outcomes
- Integration with established prognostic markers (EGFR, IDH)
- Alignment with SPIRIT guidelines for transparent protocol reporting

#### Limitations

- Non-randomized, observational design limits causal inference; residual confounding possible
- Single biomarker assessment without multi-omics integration (transcriptomics, metabolomics)
- IHC variability despite standardization; no central pathology review by external laboratory
- Potential selection bias toward surgical candidates (KPS ≥70 requirement)
- Limited sample size may restrict subgroup analyses and interaction testing
- Follow-up limited to 24 months; late recurrences and survival patterns not captured
- Generalizability limited to Colombian public healthcare system; may not apply to other settings
- The study does not include systematic assessment of chromosome 7/10 copy number status or EGFR amplification by FISH, which may limit interpretation of PINK1-EGFR interactions. Future studies should incorporate comprehensive cytogenetic profiling to elucidate these relationships.

## Data Availability

Data sharing is not applicable to this article as no datasets were generated or analysed during the current study. This is a protocol for a observational study that has not yet initiated recruitment. The deidentified dataset and statistical code will be made available upon reasonable request to the corresponding author following study completion, in accordance with FAIR principles.

## Author Contributions

Luis Alejandro García Rairán, MD Principal Investigator; protocol design; patient recruitment; surgical coordination; data collection; analysis; manuscript drafting Resident in Neurosurgery, Universidad Nacional de Colombia; Hospital de Kennedy Andrés David Turizo Smith, PhD Co-investigator; scientific direction; biomarker analysis supervision; methodology design; manuscript review. Center for Research and Training in Neurosurgery (CIEN), Samaritana University Hospital, Bogotá, Colombia. Grupo de investigación en Fitoquímica y Farmacognosia (GIFFUN), Universidad Nacional de Colombia,Bogotá 111221, Colombia.

Valentina Corpus Gutiérrez, MD, Clinical research fellow; patient follow-up support; clinical data collection; database curation; assistance in manuscript preparation. Center for Research and Training in Neurosurgery (CIEN), Samaritana University Hospital, Bogotá, Colombia.

Maria A. Del Castillo, MD, Clinical research fellow; patient screening; clinical documentation support; data verification; assistance in literature review and manuscript preparation. Center for Research and Training in Neurosurgery (CIEN), Samaritana University Hospital, Bogotá, Colombia.

William Riveros Castillo, MD, Senior neurosurgeon; clinical case review; surgical case advisory; institutional clinical support; manuscript review. Center for Research and Training in Neurosurgery (CIEN), Samaritana University Hospital, Bogotá, Colombia.

Javier Saavedra Gerena, MD, Senior neurosurgeon; clinical oversight; complex case discussion; surgical advisory; manuscript review. Center for Research and Training in Neurosurgery (CIEN), Samaritana University Hospital, Bogotá, Colombia.

Lorena Regina Rengifo, MD-Pathologist; IHC protocol development; tissue processing; blinded scoring; pathology review Subred Sur Occidente Pathology Service.

Jaime Andelfo Arias Guatibonza, MD, Senior neurosurgeon Co-investigator; surgical oversight; institutional coordination; mentorship; manuscript review, Coordinator, Neurosurgery Program, Universidad Nacional de Colombia; Hospital de Kennedy.

## Patient And Public Involvement

Patients and the public were not involved in the design of this study. Results will be disseminated to participants and their families through written summaries upon request. A patient representative will be invited to join the trial steering committee for the results interpretation phase.

## Trial Status

Protocol version: 2.0 (January 27, 2026)

Recruitment start date: August 2025 (anticipated)

Recruitment end date: December 2027

Last follow-up: October 2028

